# The research trends and hotspots of NLRP3 inflammasome in Alzheimer’s disease: a bibliometric and visualization analysis

**DOI:** 10.1101/2025.01.08.25320239

**Authors:** Xuanwei Wen, Huiye Yang, Shuangxi Chen, Zijian Xiao

## Abstract

**Background:** Alzheimer’s disease (AD) is the most common neurodegenerative disorder, the NLRP3 inflammasome has been shown to play a pivotal role in the pathogenesis of AD, and with increasing attention to its involvement in AD. Therefore, we applied bibliometric methods to describe the current research status of the NLRP3 inflammasome in AD. This study aims to analyze the research trends and hotspots in this field from 2013 to 2024, providing valuable insights for AD research.

**Methods:** We have selected research on the NLRP3 inflammasome in Alzheimer’s disease from the Web of Science Core Collection, with the time range from January 1, 2013, to November 30, 2024, and exported all publications in plain text format. Visualization analysis was performed using CiteSpace 6.4.R1, VOSviewer 1.6.20, and Scimago Graphica 1.0.46.

**Results:** A total of 759 publications related to the NLRP3 inflammasome in Alzheimer’s disease were included in this study. The number of annual publications showed a general upward trend. The top three countries in terms of publication volume were China, the United States, and Italy. The University of Manchester was the institution with the highest number of publications. The author with the most publications was Michael Heneka, while the most cited author was Eicke Latz. The International Journal of Molecular Sciences published the highest number of articles and was also the most frequently cited journal. The most common keywords included Alzheimer’s disease, NLRP3 inflammasome, neuroinflammation, Aβ, and microglia.

**Conclusion:** The primary research hotspots in this field focus on the role of NLRP3 in AD pathology, its potential as a therapeutic target, and strategies to modulate neuroinflammation through targeting the NLRP3 inflammasome. Future research should further investigate the interactions between NLRP3 and other molecular pathways, assess its clinical therapeutic potential, and provide new insights and strategies for the early diagnosis and treatment of AD.

## 1. Introduction

Alzheimer’s disease (AD) is a common neurodegenerative disorder among the elderly, primarily characterized by progressive cognitive dysfunction and behavioral abnormalities(1). With the aging global population, AD has become a significant public health challenge, affecting millions of individuals and continuing to grow rapidly (2). It is projected that by 2050, the global prevalence of AD will reach approximately 152 million, with a particularly high growth rate in developing countries(3). The pathogenesis of AD is complex and multifactorial, involving genetic and environmental factors(4), Aβ toxicity(5), tau hyperphosphorylation(6), oxidative stress(7), cholinergic changes(8), mitochondrial dysfunction(9), and endoplasmic reticulum stress(10), among others. Despite significant advances in AD research, the complete elucidation of its pathological mechanisms remains elusive. Given the extensive global impact of AD, identifying effective therapeutic targets and developing targeted therapies is of paramount importance.

Numerous studies have demonstrated that neuroinflammation plays a central role in the onset and progression of AD(11–13), with increasing attention on the role of the NLRP3 inflammasome in AD(14). The NLRP3 inflammasome is a multi-protein complex considered a key regulator of the innate immune response, playing a central role in the activation of immune responses(15). Its major components include cytoplasmic pattern recognition receptors (PRRs), the adaptor protein ASC, and the effector enzyme caspase-1(16). Under various danger signals, the NLRP3 inflammasome is activated, leading to the cleavage of inflammatory cytokines such as IL-1β and IL-18, thus triggering an inflammatory response(17). In recent years, research on the role of the NLRP3 inflammasome in Alzheimer’s disease has significantly increased, with many studies focused on its potential as a therapeutic target(18–20). The development of NLRP3 inhibitors has particularly attracted widespread attention, as these molecules may represent a novel strategy to alleviate inflammation and attenuate the progression of AD(21, 22). Existing studies have shown that inhibition of the NLRP3 inflammasome can reduce amyloid plaque deposition(23), inhibit tau hyperphosphorylation(24), and improve cognitive deficits in AD animal models(25). However, despite promising results from preclinical animal studies, targeted therapy of the NLRP3 inflammasome in clinical trials may yield inconsistent outcomes due to drug toxicity(26), highlighting the complexity of targeting neuroinflammation in Alzheimer’s disease. This discrepancy underscores the need for a comprehensive understanding of the molecular mechanisms of the NLRP3 inflammasome in Alzheimer’s disease.

Bibliometric analysis has become an essential tool for tracking scientific research trends and hotspots(27). By systematically analyzing published literature, it highlights research output, identifies influential studies and collaboration patterns, and provides insights into emerging research areas. To gain a deeper understanding of the contribution of the NLRP3 inflammasome in AD research, we conducted a bibliometric analysis of the relevant literature in this field. A statistical and visualization analysis was performed on studies published between 2013 and 2024, providing a comprehensive review and analysis of research trends and hotspots in this area. This study aims to offer valuable references and ideas for researchers in the field, further deepen the understanding of the role of the NLRP3 inflammasome in AD, and lay the foundation for the development of innovative therapeutic strategies.

## 2. Material and Methods

### 2.1 Data source

The bibliometric data was sourced from the Web of Science Core Collection (WOSCC), which is the preferred database for bibliometric research(28). The detailed selection process is illustrated in Fig 1.

**Fig 1.**
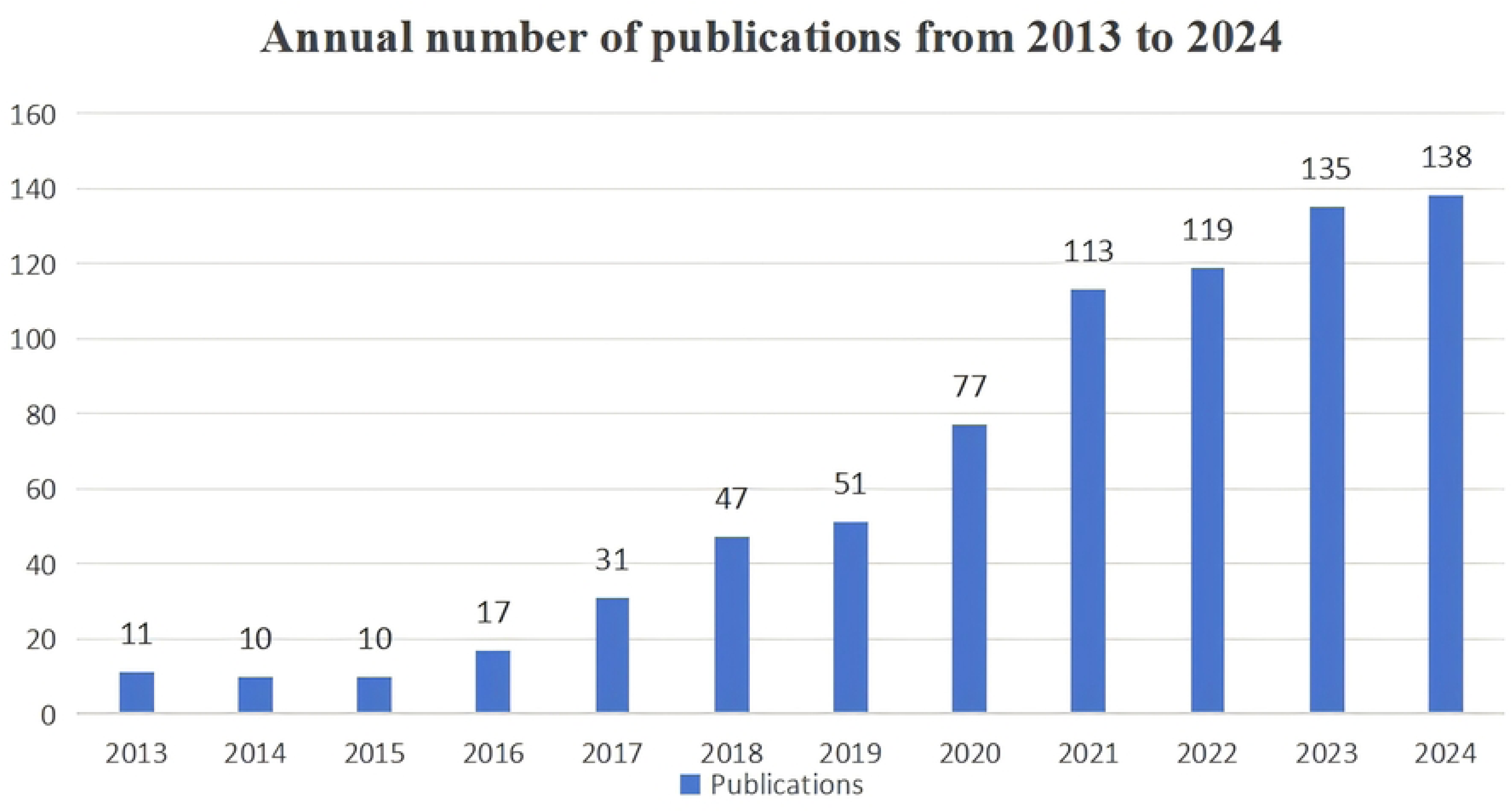
The search and filtering process.

### 2.2 Search strategy

To ensure a comprehensive and precise literature search, the following retrieval strategy was employed in the WOSCC database: TS = (“NLRP3 inflammasome” OR “NLRP3”) AND TS = (“Alzheimer’s Diseases” OR “Alzheimer Diseases” OR “Alzheimers Diseases” OR “Alzheimer Dementia” OR “Alzheimer Dementias” OR “Dementia, Alzheimer” OR “Dementia, Alzheimer Type” OR “Alzheimer Type Dementia” OR “Alzheimer Type Senile Dementia” OR “Alzheimer Sclerosis” OR “Sclerosis, Alzheimer” OR “Alzheimer Syndrome” OR “Senile Dementia, Alzheimer Type” OR “Alzheimer’s Disease” OR “Alzheimer Disease, Late Onset” OR “Late Onset Alzheimer Disease” OR “Alzheimer’s Disease, Focal Onset” OR “Focal Onset Alzheimer’s Disease” OR “Alzheimer Disease, Familial (FAD)” OR “Familial Alzheimer Disease (FAD)” OR “Familial Alzheimer Diseases (FAD)” OR “Alzheimer Disease, Early Onset” OR “Early Onset Alzheimer Disease” OR “Presenile Alzheimer Dementia” OR “Alzheimer-Type Dementia (ATD)” OR “Alzheimer Type Dementia (ATD)” OR “Dementia, Alzheimer-Type (ATD)”). The time frame for the search was limited to publications from January 1, 2013, to November 30, 2024.

### 2.3 Data collection and analysis

The authors systematically collected literature from the WOSCC database based on the following criteria: (1) the topic and content of the literature are related to NLRP3 inflammasome and AD; (2) the article types include reviews and research articles; (3) the language of the article is English. After applying these criteria, a total of 759 articles were selected, and the full-text records were exported in plain text format. We utilized VOSviewer 1.6.20, CiteSpace 6.4.R1, and Scimago Graphica 1.0.46 for bibliometric statistics and visualization analysis to gain a clearer understanding of the development trends and cutting-edge topics in this field. Descriptive statistical analysis of parameters such as authors, countries, journals, institutions, and keywords was performed using Excel. Additionally, we conducted visualization analysis of authors, co-cited authors, countries, institutions, journals, and keywords using VOSviewer 1.6.20. In the visualizations, the size of the nodes is proportional to their frequency of occurrence, and the connections between nodes represent collaboration or relationships. CiteSpace, developed by Chaomei Chen, is a Java-based application designed for the visualization and analysis of evolutionary patterns in published literature(29). Typically, each node represents an analytical factor, and the size of the node corresponds to the frequency of the data. The higher the frequency, the larger the node. Connections between nodes represent interrelationships, collaborations, or co-citation relationships. We primarily used CiteSpace 6.4.R1 to analyze the relationships between source journals and cited journals, focusing on research domains, disciplines, and the dynamic changes in emergent keywords. At the same time, CiteSpace 6.4.R1 was used to detect duplicate literature and to analyze the annual publication volume. Scimago Graphica 1.0.46 was employed to create geographical maps that display the publication output and collaboration networks of publishers by country.

Price’s Law states that in a specific field, the number of core contributors (researchers or institutions) is approximately the square root of the total number of researchers or institutions. Through further derivation, the formula is obtained as: M=0.749× √n_max_, Where M represents the minimum publication count of core authors, and n_max_ refers to the total number of publications by the highest-producing author in the field. Authors whose publication count exceeds M are considered core contributors. This formula is used to identify core authors, co-cited authors, countries, institutions, and keywords in the field.

## 3. Result

### 3.1 Analysis of annual publications

We selected 759 articles related to NLRP3 inflammasomes and AD published between 2013 and 2024 from WOSCC. After sorting the data by year (Fig 2), we observed a general upward trend in research on NLRP3 and AD since 2013. From 2013 to 2016, the growth in research output was modest, indicating that the field was still in a bottleneck phase. However, starting in 2017, the annual publication volume increased significantly and has continued to rise gradually. Notably, from 2020 to 2024, research on NLRP3 and AD surged dramatically, with the most significant increase occurring between 2020 and 2021. This trend indicates that an increasing number of scholars are interested in exploring NLRP3 inflammasomes as a potential therapeutic strategy for AD, and NLRP3 has become a hot topic in AD research.

**Fig. 2.**
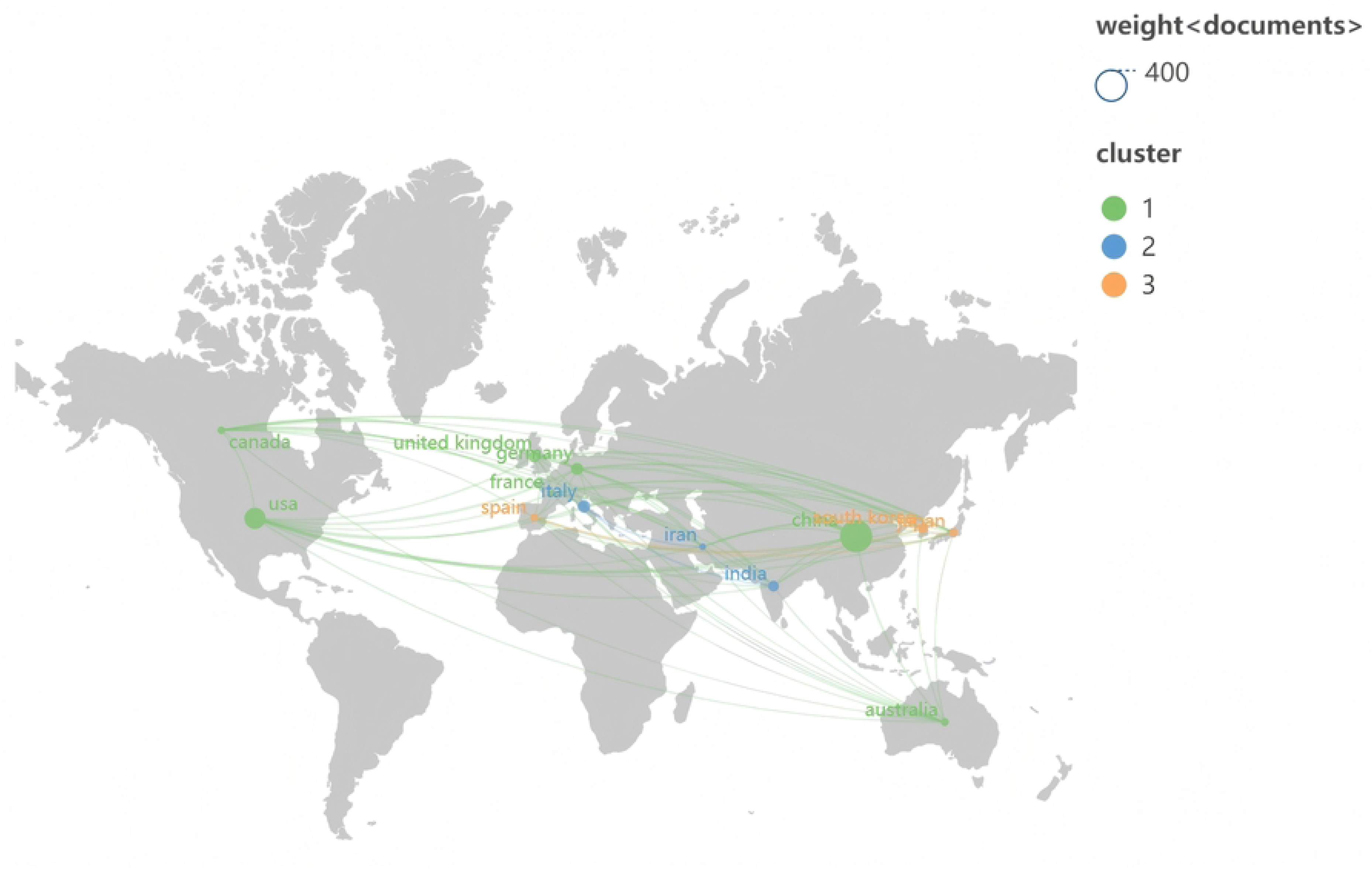
Annual Publication Volume Statistics from 2013 to 2024.

### 3.2 Analysis of countries

Between 2013 and 2024, a total of 63 countries conducted research on NLRP3 inflammasomes and their relationship with AD. The Table 1 shows the top ten countries by publication volume, with China leading the field with 351 publications, accounting for 46.25% of the total. The United States ranks second with 154 articles, making up 20.29% of the total. Together, these two countries contribute 66.54% of the total publications, indicating their dominant position in this research area. In terms of total citations, the United States holds the top spot with 17,578 citations. When considering the average citations per article, Germany ranks first with 168.76 citations per article.According to Price’s Law, countries with more than 14 publications are defined as core publishing nations. A cooperation network map of these countries was created (Fig 3). In this network, the node size corresponds to the publication volume, and the lines between nodes represent collaborative relationships, with the thickness of the lines indicating the degree of collaboration. The collaboration among core publishing countries is largely centered around the United States, which has the highest centrality of 0.40. This highlights the significant role that the United States plays in international cooperation, emphasizing its major contributions to the research on NLRP3 inflammasomes in AD.

**Fig. 3.**
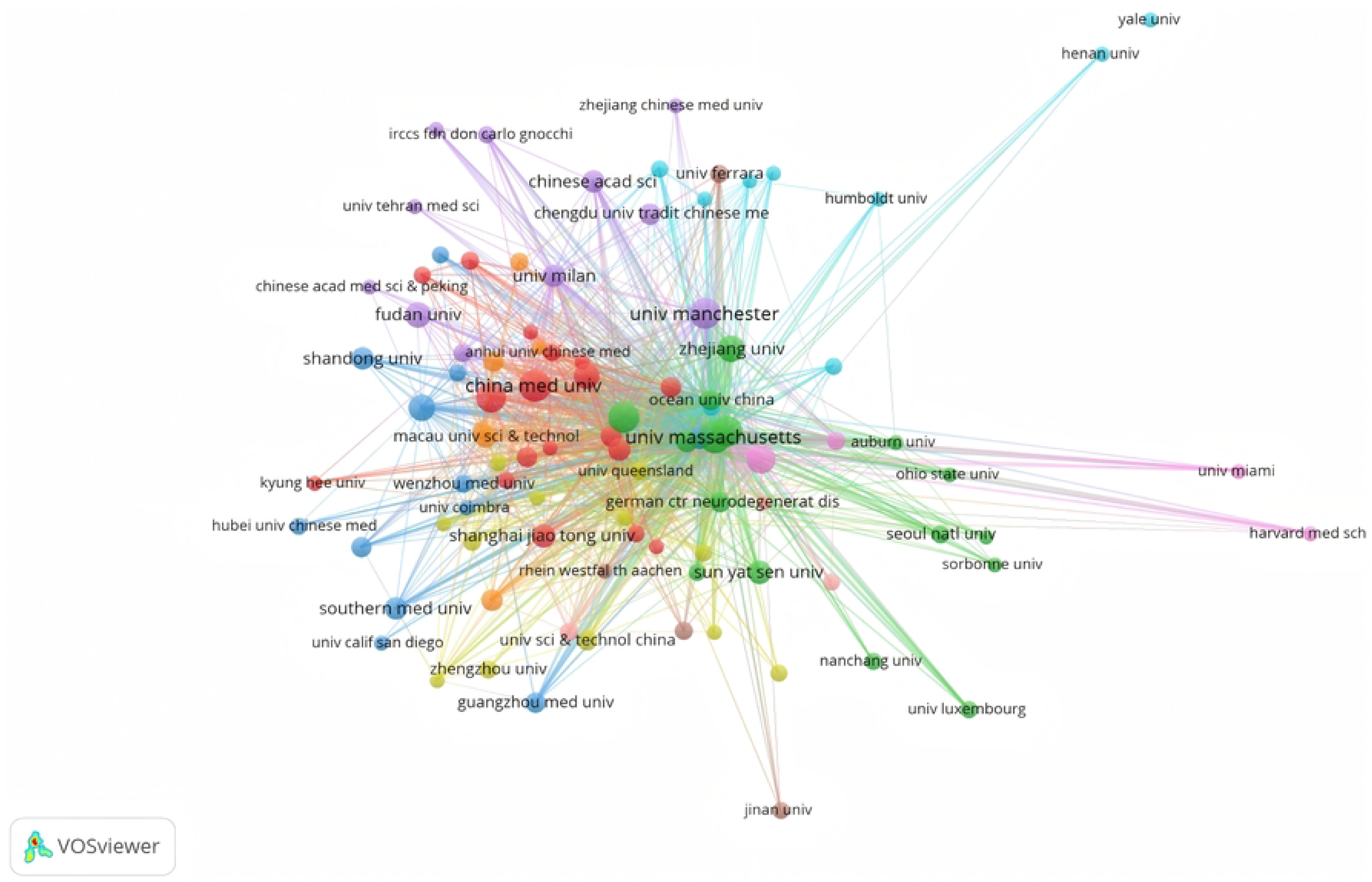
The geographical visualization map of collaboration among core countries. Each circle represents a country, with the size of the circle proportional to the number of publications from that country. Larger circles indicate a higher publication volume. The lines connecting the circles represent the collaborative relationships between countries, with the thickness of the lines reflecting the level of collaboration, thicker lines indicate closer cooperation. Additionally, lines in different colors represent distinct collaboration clusters.

**Table 1.** TOP 10 most productive countries. TP=Total Publications; TC=Total Citations

| Rank | Country | TP | TC | TC/TP | Centrality | percentage |
| --- | --- | --- | --- | --- | --- | --- |
| 1 | China | 351 | 11388 | 32.44 | 0.14 | 46.25% |
| 2 | USA | 154 | 17578 | 114.14 | 0.40 | 20.29% |
| 3 | Italy | 52 | 3111 | 59.83 | 0.14 | 6.85% |
| 4 | Germany | 51 | 8607 | 168.76 | 0.24 | 6.72% |
| 5 | England | 45 | 3024 | 67.20 | 0.27 | 5.93% |
| 6 | India | 40 | 902 | 22.55 | 0.15 | 5.27% |
| 7 | Japan | 26 | 922 | 35.46 | 0.06 | 3.43% |
| 8 | Australia | 22 | 3043 | 138.32 | 0.12 | 2.90% |
| 9 | Spain | 22 | 3389 | 154.05 | 0.09 | 2.90% |
| 10 | Canada | 20 | 954 | 47.70 | 0.02 | 2.64% |

### 3.3 Analysis of major institutions

Between 2013 and 2024, a total of 1,073 institutions were involved in research on NLRP3 inflammasomes in AD. The Table 2 provides information on the top ten institutions by publication volume. The University of Massachusetts ranks first with 19 publications, followed by China Medical University with 18 publications, and the University of Bonn with 17 publications.In terms of citation counts among the top ten publishing institutions, the University of Massachusetts received a total of 6,957 citations, making it the most cited institution. It also ranks first in terms of average citations per article, with 366.16 citations per publication. According to Price’s Law, institutions with more than 4 publications are considered core publishing institutions, with 95 institutions falling into this category(Fig 4).Although the University of Manchester published fewer papers, it ranks first in centrality with a score of 0.12, indicating strong collaboration with other institutions and a central position in the research network. The University of Bonn, Nanjing Medical University, and the German Center for Neurodegenerative Diseases follow closely with a centrality score of 0.07 each, highlighting their significant collaborative role within the field.

**Fig 4.**
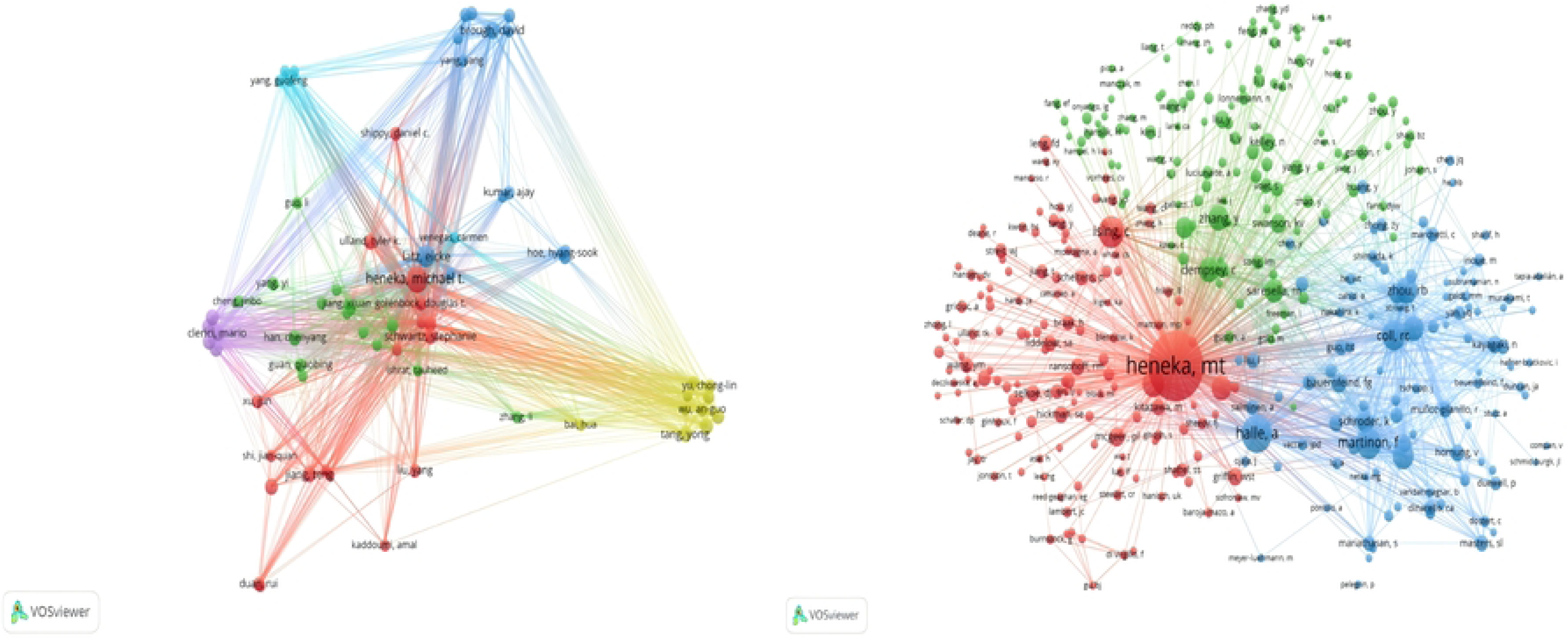
The collaboration network map among core institutions. Each circle represents an institution, with the size of the circle proportional to the number of publications from that institution. Larger circles indicate a higher publication volume. The lines connecting the circles represent the collaborative relationships between institutions, with the thickness of the lines reflecting the strength of the collaboration, thicker lines denote closer cooperation. Additionally, lines in different colors represent distinct collaborative clusters.

**Table 2.** TOP 10 most productive institutions.

| Rank | institutions | TP | TC | TC/TP | Centralit<br>y |
| --- | --- | --- | --- | --- | --- |
| 1 | University of Massachusetts | 19 | 6957 | 366.16 | 0.01 |
| 2 | China Medical University | 18 | 865 | 48.056 | 0.02 |
| 3 | University of Bonn | 17 | 5918 | 348.12 | 0.07 |
| 4 | Nanjing Medical University | 17 | 975 | 57.35 | 0.07 |
| 5 | University of Manchester | 17 | 892 | 52.47 | 0.12 |
| 6 | China Pharmaceutical University | 16 | 340 | 21.25 | 0.02 |
| 7 | German Center for Neurodegenerative Diseases | 14 | 1592 | 113.71 | 0.07 |
| 8 | Capital Medical University | 12 | 304 | 25.33 | 0.02 |
| 9 | Zhejiang University | 12 | 1406 | 117.17 | 0.03 |
| 10 | Fudan University | 11 | 112 | 10.18 | 0.02 |

TP=Total Publications; TC=Total Citations

### 3.4 Authors and co-cited authors analysis

Between 2013 and 2024, a total of 4,457 authors participated in research on NLRP3 inflammasomes in AD. The Table 3 presents information on the top ten authors ranked by publication volume. The most prolific author is Michael Heneka from the University of Bonn, followed by Eicke Latz, also from the University of Bonn, and David Brough from the University of Manchester.The fact that the top two authors, Michael Heneka and Eicke Latz, are both affiliated with the University of Bonn indicates that this institution plays a leading role in this research area. Notably, the publication volumes of the top ten authors show only small differences, suggesting that the field still holds significant potential for further exploration. Additionally, none of these top authors exhibit high centrality, indicating that their collaborations are relatively dispersed, and there isn’t a tightly concentrated research network among them.According to Price’s Law, authors with four or more publications in the field are considered core authors, with 68 core authors identified. The visualization of their collaborative network shows the relationships among these core contributors(Fig 5A).In the citation frequency analysis, co-cited authors are those who are cited together in the same reference list. In the co-citation network of NLRP3 inflammasomes and AD, there are 31,751 co-cited authors. By limiting the analysis to authors with more than 20 citations, we identified 373 authors in the co-citation network (Fig 5B). Based on the co-citation frequency, the top three most co-cited authors are Michael Heneka (730 citations), Halle Annett (266 citations), and Ising Christina (201 citations). Fabio Martinon has the highest centrality score of 0.05 among all authors, indicating his prominent role in the network.

**Fig. 5.**
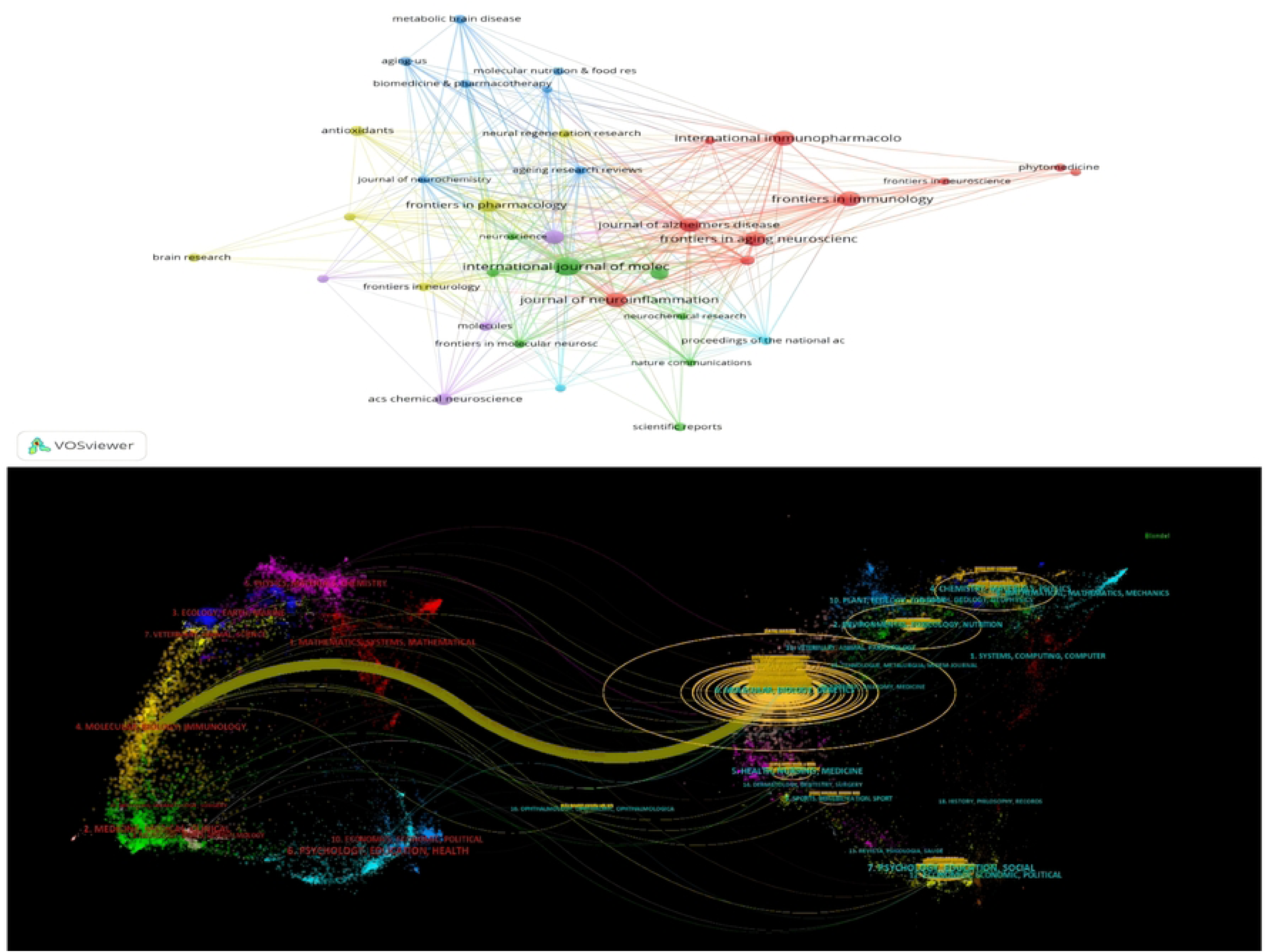
(A) The collaboration network map of core authors. (B) The collaboration network map of core co-citation authors. Each circle represents an author, with the size of the circle proportional to the number of publications by the author. The lines connecting the circles represent collaborations between authors, with thicker lines indicating closer cooperation. Different colors represent different collaboration groups of authors.

**Table 3.** TOP 10 most productive authors.

| Rank | Authors | TP | TC | TC/TP | Centrality |
| --- | --- | --- | --- | --- | --- |
| 1 | Michael Thomas Heneka | 19 | 4584 | 241.26 | 0 |
| 2 | Eicke Latz | 13 | 6842 | 526.31 | 0.01 |
| 3 | David Brough | 8 | 818 | 102.25 | 0 |
| 4 | Yu, Lu | 7 | 254 | 36.29 | 0 |
| 5 | Stephanie Schwartz | 7 | 3663 | 523.29 | 0 |
| 6 | Tang, Yong | 7 | 288 | 41.14 | 0 |
| 7 | Zhang, Ying-Dong | 7 | 259 | 37.00 | 0 |
| 8 | Marina Saresella | 7 | 516 | 73.71 | 0 |
| 9 | Jiang, Teng | 7 | 443 | 63.29 | 0 |
| 10 | Mario Clerici | 7 | 516 | 73.71 | 0 |

TP=Total Publications; TC=Total Citations

### 3.5 Journals distribution analysis

We conducted a comprehensive analysis of 289 journals exploring the relationship between NLRP3 inflammasomes and AD. These journals are predominantly published in Switzerland and the United States. As shown in the Table 4, the top three journals by publication volume are the International Journal of Molecular Sciences, Frontiers in Immunology, and Frontiers in Aging Neuroscience, with 32, 22, and 21 papers published, respectively. Notably, these journals are also the most frequently cited, indicating that they are highly valuable references in the field of NLRP3 inflammasomes and AD research.Among the top ten journals by publication volume, the Journal of Neuroinflammation has the highest impact factor at 9.3. The International Journal of Molecular Sciences has the highest citation frequency per article, with an average of 99.44 citations per paper.According to Price’s Law, journals with five or more articles are considered core journals in this field. A total of 37 journals have been identified as core journals. The visual network map helps us better understand the relationships between these core journals (Fig 6A).Furthermore, we used CiteSpace’s dual-map overlay tool, which is an effective method for revealing subject relationships within the citation network. The dual-map overlay we constructed (Fig 6B) presents a major pathway: the orange path indicates that journals related to molecular biology and genetics are cited by journals in molecular biology and immunology. This overlay reflects the cross-disciplinary citation relationships and highlights the collaboration and connection between different research fields.

**Fig. 6.**
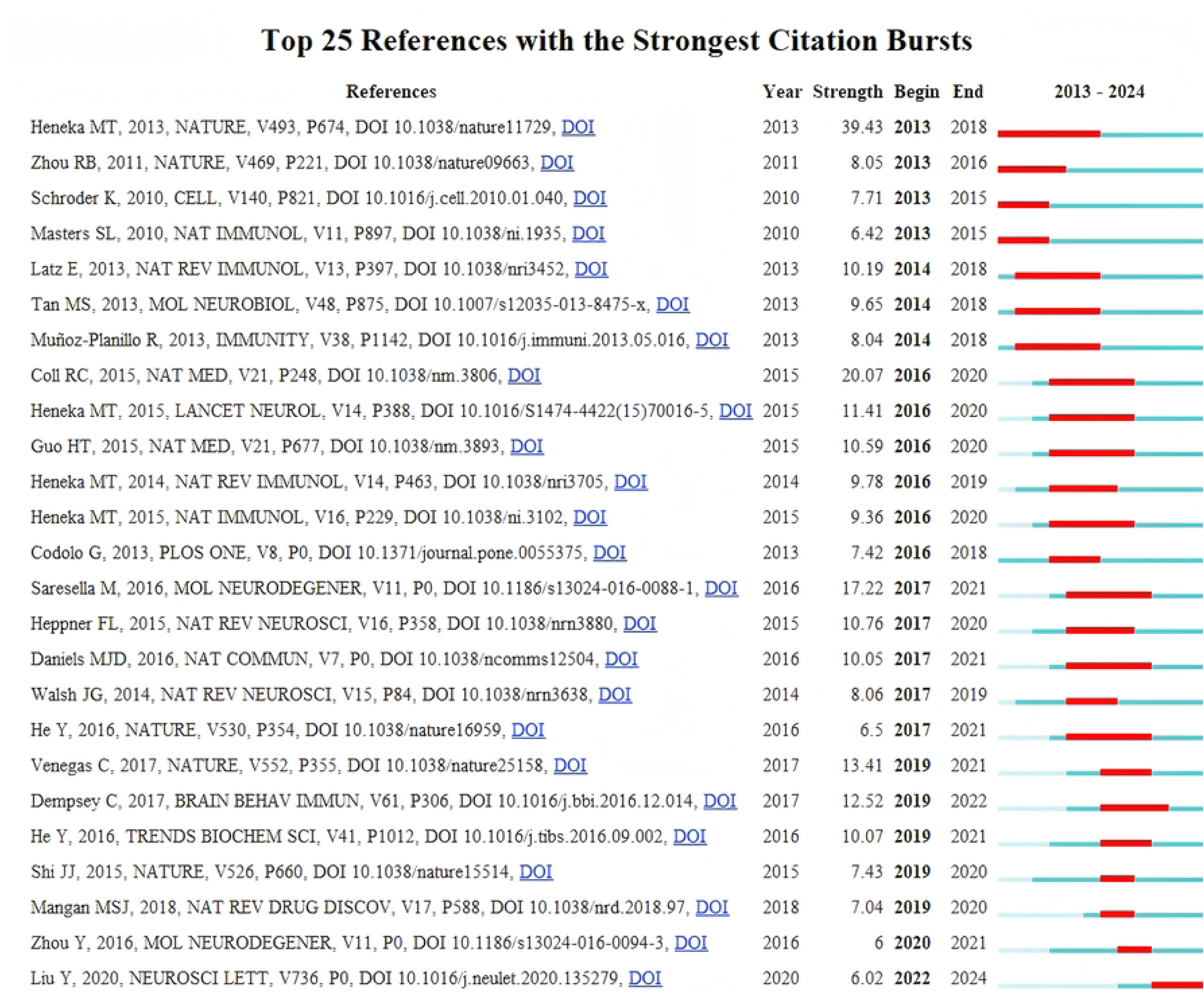
(A) The core journal visualization network map. Each circle represents a journal, with the size of the circle proportional to the number of publications. The lines connecting the circles represent the relationships between journals, with thicker lines indicating stronger connections. Different colors represent distinct journal clusters. **(B) The dual-map overlay of journals publishing research.** The left side represents the citing journals, while the right side represents the cited journals. The lines indicate citation paths, and labels in different colors represent different academic disciplines.

**Table 4.** TOP 10 most productive journals.

| Rank | journal | TP | TC | TC/TP | IF | Country |
| --- | --- | --- | --- | --- | --- | --- |
| 1 | International Journal of Molecular Sciences | 32 | 3182 | 99.44 | 4.9 | USA |
| 2 | Frontiers in Immunology | 22 | 1908 | 86.73 | 5.7 | Switzerland |
| 3 | Frontiers in Aging Neuroscience | 21 | 749 | 35.67 | 4.1 | Switzerland |
| 4 | Journal of Neuroinflammation | 20 | 691 | 34.55 | 9.3 | England |
| 5 | International Immunopharmacology | 20 | 534 | 26.70 | 4.8 | Netherlands |
| 6 | Journal of Alzheimers Disease | 19 | 448 | 23.58 | 3.4 | Netherlands |
| 7 | Molecular Neurobiology | 19 | 613 | 32.26 | 4.6 | USA |
| 8 | Frontiers in Pharmacology | 17 | 363 | 21.35 | 4.4 | Switzerland |
| 9 | Cells | 15 | 408 | 27.20 | 5.1 | Switzerland |
| 10 | ACS Chemical Neuroscience | 13 | 231 | 17.77 | 4.1 | USA |

TP=Total Publications; TC=Total Citations; IF=Impact Factor

### 3.6 Analysis of co-cited references and reference burst

We analyzed the top ten most-cited references, as shown in the Table 5. Notably, the paper “NLRP3 is activated in Alzheimer’s disease and contributes to pathology in APP/PS1 mice,” authored by Michael T. Heneka et al. in 2013 and published in Nature, is the most cited among these, with a citation count of 393. It is followed by the paper “The NALP3 inflammasome is involved in the innate immune response to amyloid-beta” by Annett Halle et al., published in Nature Immunology in 2008, which has been cited 266 times. Additionally, seven other publications have been cited over 100 times. The co-cited references in the top ten primarily focus on the activation of the NLRP3 inflammasome in AD and its pathological roles.The citation burst analysis of the references reveals the ability to identify periods of sudden citation increases for specific papers or topics, thereby highlighting the themes and fields that the academic community has focused on at different stages. As shown in the Fig 7, Michael T. Heneka’s study in 2013 exhibited the highest citation burst intensity. This study revealed that the NLRP3 inflammasome is activated in AD mice in response to amyloid-beta (Aβ), and it further contributes to the progression of AD. Moreover, the activation of the NLRP3 inflammasome and the subsequent release of inflammatory mediators were found to be associated with cognitive dysfunction, synaptic impairments, and the limited clearance function of microglia. This suggests that targeting the NLRP3 inflammasome or its downstream cytokines could be a promising therapeutic strategy for preventing the progression of AD. Recent citation trends in the referenced literature indicate a growing focus on the regulation of the NLRP3 inflammasome as a therapeutic approach for various diseases, underscoring the broad impact of the NLRP3 inflammasome. The primary therapeutic strategy involves the use of specific NLRP3 inflammasome inhibitors to modulate inflammation and immune responses, thereby achieving disease-modulating effects, such as delaying or controlling disease progression.

**Fig 7.**
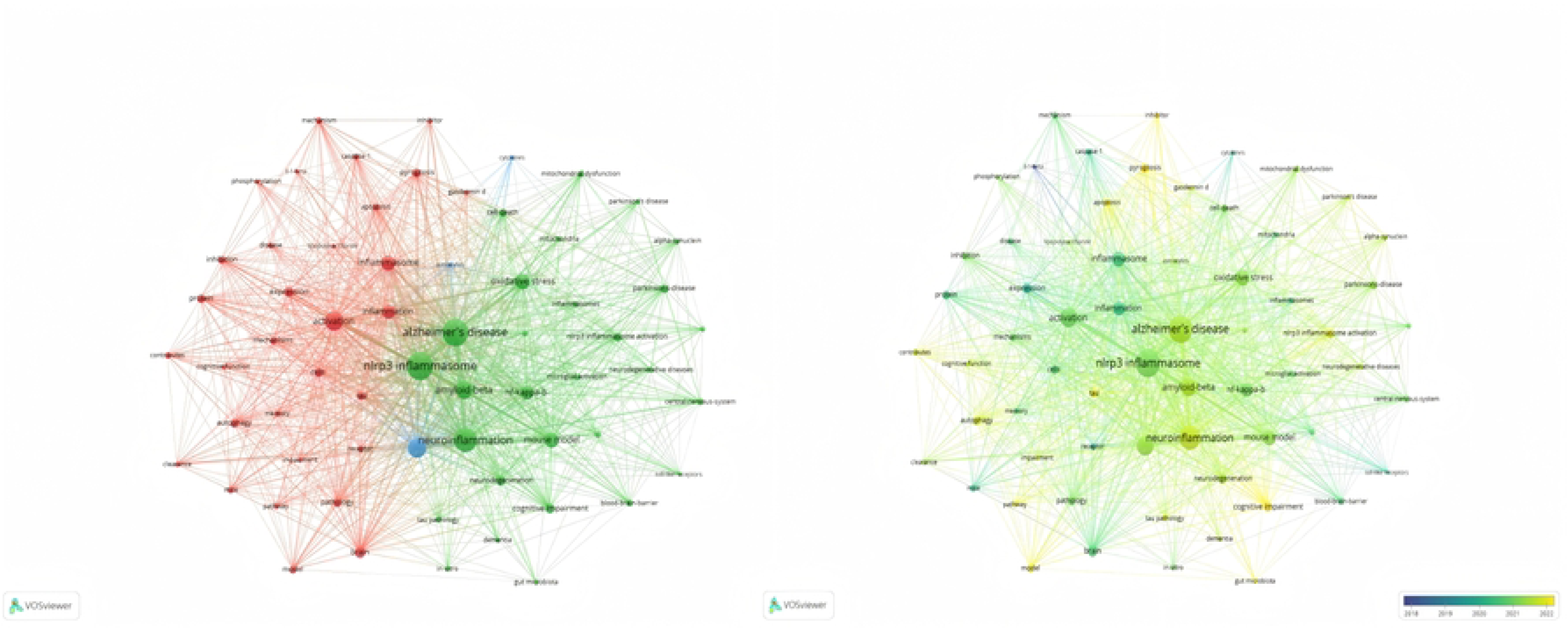
The top 25 references with the strongest bursts related to NLRP3 inflammasome in AD

**Table 5.** Top 10 most co-cited references.

| Rank | Author | References | Year | TC |
| --- | --- | --- | --- | --- |
| 1 | Michael T Heneka | NLRP3 is activated in Alzheimer's disease and contributes to pathology in APP/PS1 mice | 2013 | 393 |
| 2 | Annett Halle | The NALP3 inflammasome is involved in the innate immune response to amyloid-beta | 2008 | 266 |
| 3 | Christina Ising | NLRP3 inflammasome activation drives tau pathology | 2019 | 190 |
| 4 | C Dempsey | Inhibiting the NLRP3 inflammasome with MCC950 promotes non-phlogistic clearance of amyloid-β and cognitive function in APP/PS1 mice | 2017 | 131 |
| 5 | Carmen Venegas | Microglia-derived ASC specks cross-seed amyloid-β in Alzheimer's disease | 2017 | 122 |
| 6 | Rebecca C Coll | A small-molecule inhibitor of the NLRP3 inflammasome for the treatment of inflammatory diseases | 2015 | 116 |
| 7 | Michael T Heneka | Neuroinflammation in Alzheimer's disease | 2015 | 112 |
| 8 | Marina Saresella | The NLRP3 and NLRP1 inflammasomes are activated in Alzheimer's disease | 2015 | 111 |
| 9 | Ilie-Cosmin Stancu | Aggregated Tau activates NLRP3-ASC inflammasome exacerbating exogenously seeded and non-exogenously seeded Tau pathology in vivo | 2016 | 95 |
| 10 | Rongbin Zhou | A role for mitochondria in NLRP3 inflammasome activation | 2019 | 94 |

TC=Total Citations

### 3.7 Keyword clustering and time analysis

Keywords capture the core essence of a scientific article, and through co-occurrence analysis, they can help identify trends and hotspots within a particular field of research. As shown in the Table 6, the top three most frequent keywords in the field of NLRP3 inflammasomes and AD are “Alzheimer’s Disease,” “NLRP3 inflammasome,” and “Neuroinflammation,” with 573, 556, and 280 occurrences, respectively. According to Price’s Law, keywords that appear more than 18 times are categorized as core keywords, with a total of 69 core keywords identified. We conducted a visualization of these core keywords(Fig 8A). The size of each node is proportional to the frequency of the keyword’s appearance, while the thickness of the connecting lines represents the strength of the relationship between keywords. The different colors of the nodes represent various clusters. Cluster 1 (green) is the largest, containing 31 keywords related to activation, inflammation, and inflammasomes. Cluster 2 (red) contains 29 keywords, primarily focused on the relationship between Alzheimer’s disease, NLRP3, neuroinflammation, and Aβ. Cluster 3 (blue) focuses on microglia, astrocytes, and cytokines with three keywords. To better understand the dynamic changes in research hotspots, we used VOSviewer to generate a time distribution map for keyword clustering(Fig 8B). The color variation of the nodes reflects the time period during which the keywords emerged, with yellow nodes representing more recent trends compared to blue nodes.

**Fig. 8.**
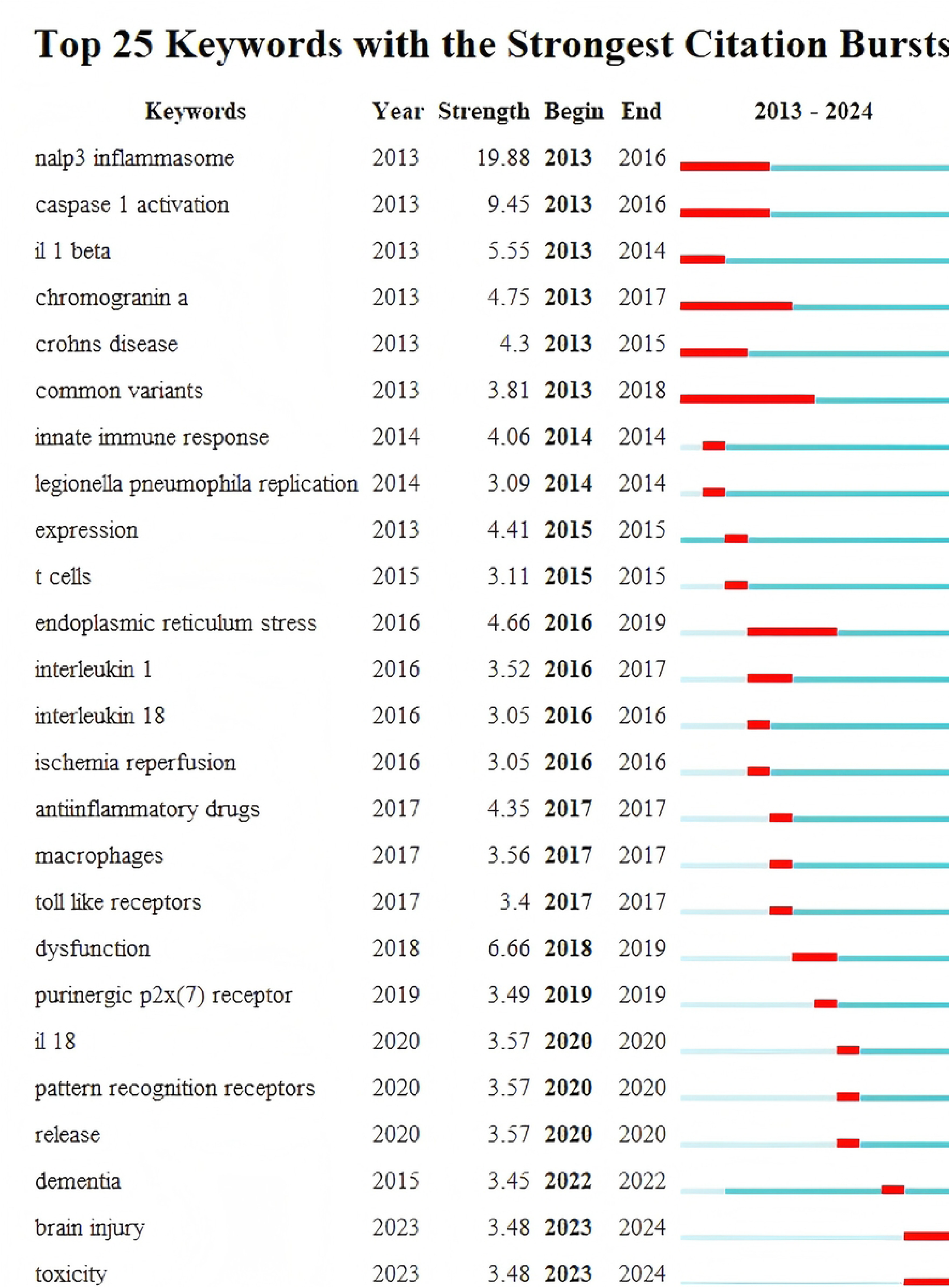
(A) The visualization network of core keyword clusters. The size of the circles is proportional to their frequency of occurrence. Different colors of the circles represent different clusters. The lines between the circles indicate co-occurrence in the same document, with the thickness of the lines representing the frequency of co-occurrence. **(B) The visualization of the temporal distribution of core keywords.** The size of the circles is proportional to their frequency of occurrence, and the color of the circles is associated with the time of the keyword’s appearance, with blue keywords appearing earlier than yellow ones.

**Table 6.** TOP 10 frenquency keywords.

| Rank | Keyword | Counts | Rank | Keyword | Counts |
| --- | --- | --- | --- | --- | --- |
| 1 | alzheimer's disease | 556 | 6 | activation | 195 |
| 2 | nlrp3 inflammasome | 506 | 7 | oxidative stress | 133 |
| 3 | neuroinflammation | 280 | 8 | mouse model | 128 |
| 4 | amyloid-beta | 244 | 9 | nf-kappa-b | 127 |
| 5 | microglia | 195 | 10 | inflammasome | 126 |

#### 3.8 Keyword burst analysis

Keyword burst analysis allows the identification of emerging trends and hot topics within a research field by analyzing the sudden increase in the frequency of keywords during a specific time period. To further explore the emerging trends and research hotspots related to the NLRP3 inflammasome in AD, we conducted a burst keyword analysis using Citespace, which identified the top 25 burst terms in this field(Fig 9). The results revealed that the term with the strongest burst intensity was “caspase 1 activation,” followed by “dysfunction” and “IL-1β.” Notably, the term with the longest burst duration was “common variants.” Other recent burst terms included “toll-like receptors,” “purinergic P2X(7),” “IL-18,” “pattern recognition receptors,” “dementia,” “brain injury,” and “toxicity,” suggesting that these themes currently represent the primary research hotspots in the study of the NLRP3 inflammasome in AD.

**Fig. 9.**
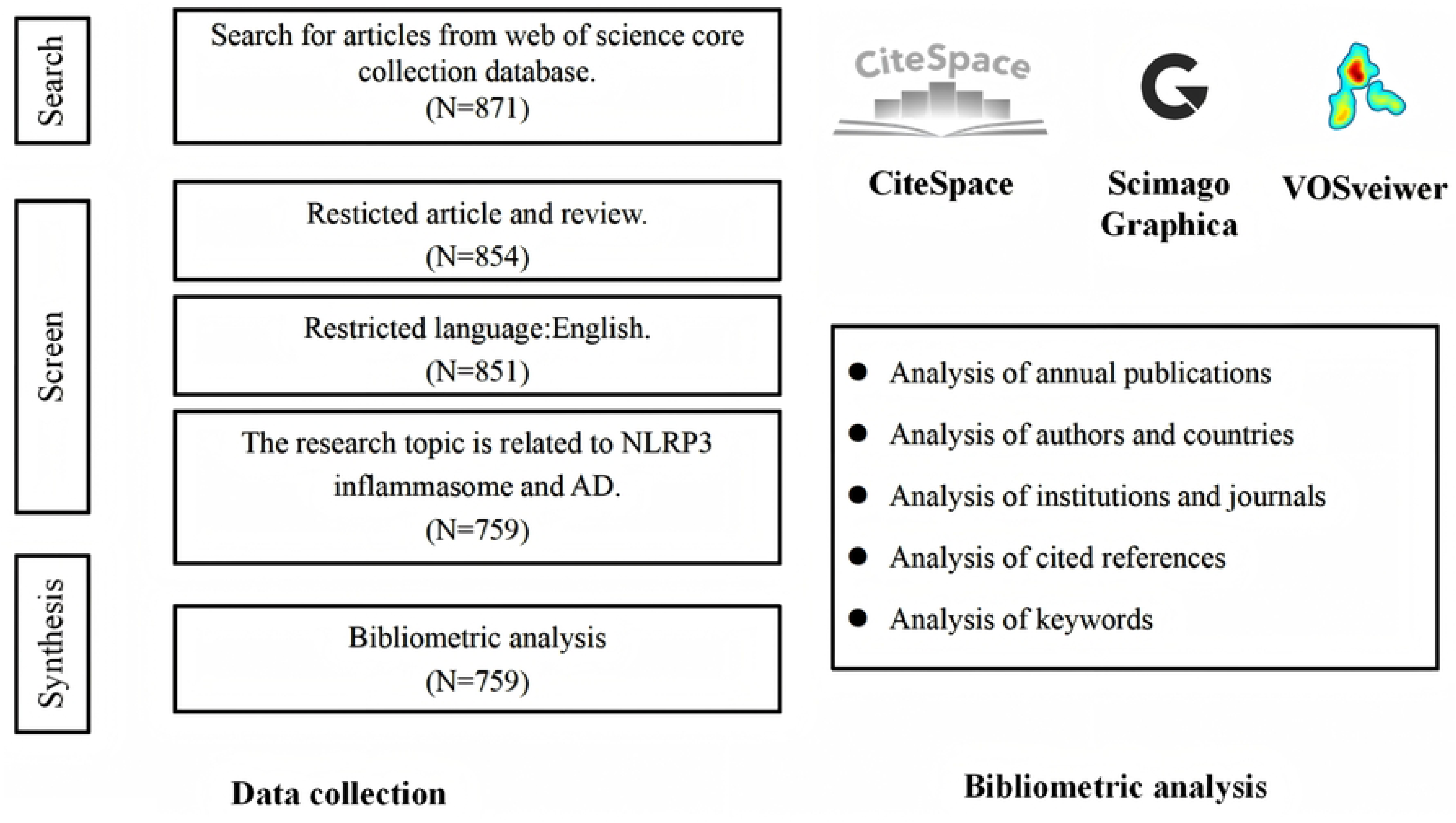
The top 25 keywords with the strongest bursts related to NLRP3 inflammasome in AD.

## 4. Discussion

### 4.1 General information

With increasing research focus on the role of the NLRP3 inflammasome in AD, it has emerged as a key factor in the pathogenesis of AD. Through bibliometric and visualization analyses, we have unveiled the research trends and hotspots of the NLRP3 inflammasome in the AD field from 2013 to 2024. This study not only provides new insights into the role of the NLRP3 inflammasome in AD but also offers important references for future research directions and therapeutic strategies. According to our analysis, since 2020, the publication volume on NLRP3 inflammasome in AD has significantly increased, indicating that the field has attracted substantial attention from scholars and entered a rapidly developing new phase.

Among all the included articles, China has emerged as the leading contributor to the global research on the NLRP3 inflammasome in AD, with 351 publications, accounting for 46.25% of the total publications. The United States ranks second, with 154 publications, representing 20.29% of the total. Although China has the highest publication volume in this field, the average citation count of its publications is relatively low, indicating that its research in this domain is not yet widely recognized and lacks substantial international impact.The collaboration network analysis shows that China has engaged in extensive cooperation with several countries, including the United States, Germany, and the United Kingdom, but the depth of these collaborations remains somewhat limited. Compared to the United States and Germany, China’s research in this area still needs to improve in terms of depth and quality. Further studies should focus on exploring more meaningful and therapeutically valuable experiments.Thus, a higher publication volume does not necessarily equate to greater contribution; quality remains the key factor. Germany, with a total of 51 publications, has the highest average citation count, reaching 168.76 citations per paper, highlighting the significant reference value of its research in this field. Notably, the two most cited studies explore the activation of the NLRP3 inflammasome in AD(14) and its involvement in Aβ-mediated immune responses, respectively(30).

Among the top ten institutions ranked by publication volume, five are from China: China Medical University, Nanjing Medical University, China Pharmaceutical University, Zhejiang University, and Fudan University. This highlights the substantial resources and funding that China has dedicated to research on the NLRP3 inflammasome in AD in recent years, attracting a large number of researchers to this field. It also reflects the presence of strong research teams and well-established research facilities in these institutions. However, institutions like the University of Manchester, Bonn University, and the German Center for Neurodegenerative Diseases (DZNE) exhibit higher centrality, indicating that strengthening interdisciplinary and interfield collaborations is crucial for enhancing international influence. Academic exchanges and collaborative research among European and American countries are relatively frequent, and China still needs to engage in more communication and deeper collaborative research with other countries. Furthermore, the International Journal of Molecular Sciences, Frontiers in Immunology, and Frontiers in Aging Neuroscience are the top three journals in this field in terms of publication volume. These journals also have high citation frequencies, indicating that articles published in these journals are key references driving research in the NLRP3 inflammasome and AD.

Among the core author group in this field, the research teams led by Professors Michael Heneka, Eicke Latz, and David Brough have published the most papers, making them the central research teams in this domain. One of the significant contributions of Professor Michael Heneka is his in-depth investigation into the role of neuroinflammation in AD(31). His research has demonstrated that the activation of the NLRP3 inflammasome is closely associated with key pathological features of AD, such as Aβ plaque accumulation(14) and tau protein phosphorylation(32), thereby providing new therapeutic targets for AD. Professor Eicke Latz’s research primarily focuses on the activation of the innate immune system and its relationship with neurodegenerative diseases(33). His team has explored the immune-related aspects of the NLRP3 inflammasome in AD, revealing that its activation not only exacerbates neuroinflammation but also accelerates the progression of AD(34). Professor David Brough’s research primarily focuses on neuroimmunology, with particular emphasis on the NLRP3 inflammasome(35, 36). His findings suggest that nonsteroidal anti-inflammatory drugs (NSAIDs) can inhibit the NLRP3 inflammasome and exert neuroprotective effects in Alzheimer’s disease (AD) mouse models(37). Furthermore, his team has identified several NLRP3 inhibitors(38, 39), which have demonstrated significant neuroprotective effects in animal models. Moreover, the dual-map visualization of this field clearly illustrates the interdisciplinary nature of the research, encompassing areas such as psychology, biology, and genetics. Clinical-related disciplines such as neurology, nursing, genetics, and immunology are also involved in the cross-disciplinary research within this field. This multi-disciplinary and cross-field collaborative relationship further highlights the significance of the research on NLRP3 inflammasome in AD.

### 4.2 Hot topic and research trend

In-depth analysis of keywords is an essential part of identifying the research trends and hotspots in a particular field. In the keyword burst analysis, “caspase 1 activation” emerges as the most intense burst keyword, indicating its central role in the research focus of the field. “Common variants” exhibits the longest burst duration, suggesting it has sustained attention from researchers over time and holds significant value for further exploration. Moreover, aside from Alzheimer’s disease and NLRP3 inflammasome, “neuroinflammation” is the most frequently occurring keyword, highlighting its core position in this research area.Cluster analysis of the keywords reveals that research on NLRP3 inflammasome in AD predominantly focuses on areas such as neuroinflammation, activation and inhibition of inflammasomes, pathological changes, glial cells, and the associated mechanisms and pathways. Cluster 1 primarily investigates the relationship between inflammasome activation, inflammation, and related mechanisms and pathways. Studies suggest that the activation of NLRP3 inflammasome triggers inflammatory responses(40), exacerbates apoptosis(41), induces pyroptosis(42), and regulates autophagy(43), ultimately leading to AD progression. Therefore, in-depth research on NLRP3 inflammasome and its inflammatory pathways in AD is of significant importance.Cluster 2 focuses on the neuroinflammation, oxidative stress, and related pathological changes caused by the activation of NLRP3 inflammasome in AD. Upon activation, NLRP3 inflammasome secretes numerous inflammatory cytokines, which affect surrounding immune cells and neurons, further intensifying the neuroinflammatory response(44). Additionally, it leads to the excessive accumulation of reactive oxygen species (ROS), which not only accelerates neuronal death through cellular damage but also further activates the NLRP3 inflammasome, creating a positive feedback loop that exacerbates oxidative stress(45). Most importantly, the activation of the NLRP3 inflammasome further exacerbates the pathological features of AD(46). Therefore, inhibitors of NLRP3 inflammasome or other intervention measures may become an effective strategy to delay and alleviate AD progression(47).Cluster 3 focuses on glial cells and related cytokines, highlighting the close association between glial cells and NLRP3 inflammasome in AD. In Alzheimer’s disease, the activation of microglial cells and astrocytes is central to the neuroinflammatory response and is closely related to the activation of NLRP3 inflammasome(48). The time distribution map of keywords reveals that recent research has been increasingly centered around mechanistic studies, such as apoptosis, pyroptosis, autophagy, and pathways. This suggests that future research will likely focus on exploring the mechanisms through which NLRP3 inflammasome influences AD, representing an emerging field in the study of Alzheimer’s disease.

Numerous studies have shown that the NLRP3 inflammasome plays a critical role in the pathogenesis and progression of AD. Research has revealed that Aβ deposition is one of the earliest pathological changes in AD(49). The aggregation of Aβ directly activates the NLRP3 inflammasome in microglial cells, leading to the production and secretion of pro-inflammatory cytokines such as IL-1β and TNF-α(30), which further exacerbate neuroinflammatory responses, Aβ accumulation, and tau protein hyperphosphorylation in AD, ultimately causing neuronal damage. Moreover, studies have demonstrated that APP/PS1 mice with NLRP3 and caspase-1 gene deficiencies show significantly improved learning and memory functions compared to normal APP/PS1 mice. Furthermore, Aβ deposition in the hippocampus and cortex of these mice is markedly reduced, which may be attributed to enhanced microglial phagocytic capacity resulting from the NLRP3 and caspase-1 gene deficiency(14). Thus, in Alzheimer’s disease, the chronic deposition of Aβ not only leads to sustained activation of the NLRP3 inflammasome, but more importantly, the activated NLRP3 inflammasome exacerbates the impairment of Aβ clearance. This provides a potential avenue for intervening in the clinical symptoms and associated pathology of AD. Notably, evidence suggests that Aβ can activate the NLRP3 inflammasome, leading to abnormal tau phosphorylation and aggregation. The NLRP3 inflammasome plays a key role in the Aβ-tau cascade(32).Given the pivotal role of NLRP3 inflammasome in AD, many studies have begun to focus on its potential as a therapeutic target. For example, MCC950 has been shown to effectively inhibit the activation of the NLRP3 inflammasome, thereby reducing neuroinflammatory responses and improving cognitive performance in AD mouse models(19). Another study showed that, after treatment with BHB, the size and volume of Aβ plaques in the cerebral cortex of APP/PS1 transgenic Alzheimer’s disease mice were significantly reduced. These studies underscore the important role of NLRP3 inflammasome in the pathogenesis of AD, making it a highly valuable therapeutic target. Therefore, further exploration of the mechanisms associated with the NLRP3 inflammasome may offer new targets and intervention strategies for the early diagnosis, treatment, and prognosis of AD.This emerging research area has gradually become the focus of NLRP3 inflammasome research in AD, providing promising pathways for the prevention and treatment of AD. As such, in-depth studies of the NLRP3 inflammasome activation process and interventions at various sites to explore their therapeutic efficacy are highly significant. This represents a key research focus and future direction in this burgeoning field.

### 4.3 Strengthens and limitations

We conducted the first bibliometric analysis of research on the NLRP3 inflammasome in AD, providing an in-depth exploration of the research trends and hot topics in this field. Our study analyzes and summarizes the forefront of research from multiple dimensions. The aim of this research is to provide valuable insights and references for future research directions and scholars in this field. However, it is important to acknowledge the limitations of our investigation.First, we only included data obtained from the Web of Science Core Collection (WOSCC), which may result in an incomplete collection of publications, leading to potential conclusion biases due to the lack of comprehensive inclusion. In addition, we utilized multiple software tools, including Citespace, VOSviewer, and Scimago Graphica, for visualization analysis, which may introduce systematic errors during the software analysis process. Secondly, the literature retrieval process may be influenced by the researchers conducting the search, introducing a degree of subjectivity in the selection of articles. Finally, we must acknowledge that our search covered a limited time range, encompassing only publications from 2013 to 2024. Therefore, the 759 articles we selected may not fully represent all available literature in this field.Thus, future studies should consider a longer timeframe and a more comprehensive database to validate and expand upon our findings.

## 5. Conclusion

We screened 759 publications from the Web of Science Core Collection (WOSCC), with contributions from 63 countries, 1,073 institutions, and 4,457 authors in the research on the NLRP3 inflammasome in AD. Notably, the number of publications on the NLRP3 inflammasome in AD has gradually increased since 2013, with a significant rise in publication volume over the past four years, indicating a growing international attention to this field. As the first study to use bibliometric analysis to evaluate the research in this area, our work provides exceptional reference value for future explorations in this field and serves as a cornerstone for innovative research.

## Data Availability

All relevant data are within the manuscript and its Supporting Information files.

## Acknowledgments

Not applicable.

## Author contributions

WX and YH conceived the study, conducted the data analysis and interpretation, and wrote the manuscript. XZ and CS were responsible for reviewing and editing. All authors contributed to the article and approved the submitted version.

## Funding

This study was supported by the National Clinical Key Specialty Project (Grant no. 20230383).

## Notes

### Competing Interest Statement

The authors have declared no competing interest.

### Funding Statement

Yes

